# Dynamic treatment regime in critical ill patients with high risk of antibiotic drug resistance, a cohort study

**DOI:** 10.1101/2025.01.22.25320993

**Authors:** Yiwei Liu, Zhaohui Bai, Zhaoshuai Ji

## Abstract

Antibiotic resistance threatens modern medicine, especially in critical care. This retrospective observational cohort study, based on MIMIC–IV v2.0 database, included 2,532 ICU admissions with high - risk antibiotic resistance. We aimed to find the optimal strategy for using high - risk resisted antibiotics in critical ill patients by constructing a dynamic treatment regime (DTR) model with DWSurv method. The results showed that the DTR optimal model’s suggestion differed from clinical practice. Clinical practice showed a sharp decline in high - risk resistance antibiotic administration, while the model suggested treatment with gaps in 7 days. Survival analysis indicated that the DTR model had a lower mortality rate. The more the clinical practice disobeyed the model’s recommendation, the higher the 28 - day mortality (adjusted HR = 1.21, 95%CI: 1.12 - 1.30, P<0.001). This study suggests that the DTR model might offer a new perspective on personalized antibiotic administration, but its clinical application remains to be further explored.

## Introduction

Although antibiotics have saved many lives and enabled uncountable medical practices, their wide-spread use in human and animal populations has led to the rise of antibiotic resistance, posing now a threat to modern medicine, especially in critical care units. Traditional 7-day, single dose, aggressive treatment strategy might be harmful in this case since prolonged antibiotic exposure drives emergence of resistance and increases the occurrence of adverse effects [1]. Previous evidences tried to use shorter but fixed administration period to and most of them found out that the outcome was equal to long period ones in critical ill patients [2]. Also, antimicrobial stewardship program based on sterile sites was successful [3]. However, to our knowledge, no clinical study tried to investigate the best strategy to use resisted antibiotic, since most of the studies were trying to avoid or de-escalation using known high risk resisted antibiotics in this scenario [4, 5]. Compared to aggressive and static strategy, a study tried to discover more dynamic antibiotic stop timing in gram negative infection population according to CRP level [6], this earlier stop might reduce drug resistance in whole picture. However, none of the pathogens were drug resistance.

Some recent studies tried to discuss a new strategy called dynamic treatment regime (DTR) based on revolutionary theory in this circumstance, which not pursuing cure but control [7-9]. In brief, this strategy following these principles:

1. Maximum tolerable dose: periodic administration of a high dose, limited by toxic side effects.
2. Intermittent therapy: periodic administration of a high dose with fixed, periodic treatment holidays.
3. Adaptive therapy (dose skipping): adaptive dosing where a high dose is administered until a desired tumor/pathogen response (e.g., 50% size/density reduction), followed by a treatment holiday until a desired upper threshold (e.g., 100%) and repeated.
4. Adaptive therapy (dose modulation): adaptive dosing where dose is modulated (increased or decreased) at regular intervals depending on tumor/pathogen response. And previous evidence has proved this concept would prolong tumor control in drug resistance cancer patients [10, 11].

Hence the treatment decision had to adaptively adjusted according to pathogen density and host’s condition, which is dynamic through the whole process. The biggest difference between de-escalation and DTR was the latter one might start a few very short treatment durations in the whole process, instead of never started again once stopped. In DTR methodology perspective, the Q-learning based method has been widely conducted [12], which is a recursive learning technic to optimize intervention at each stage.

Theoretically, if the conventional therapy (de-escalation only, no re-start) is actually the optimal treatment strategy, the DTR optimal model would return a similar strategy or at least not privilege among conventional one. On the contrary, if DTR model returned a strategy with re-starting antibiotics, and this model showed significantly better survival among conventional strategy, then we may re-consider if we really have to stop permanently when we found certain antibiotic was resisted. The state-of-art of this study is shown as **Figure 1**:

**Figure 1.**
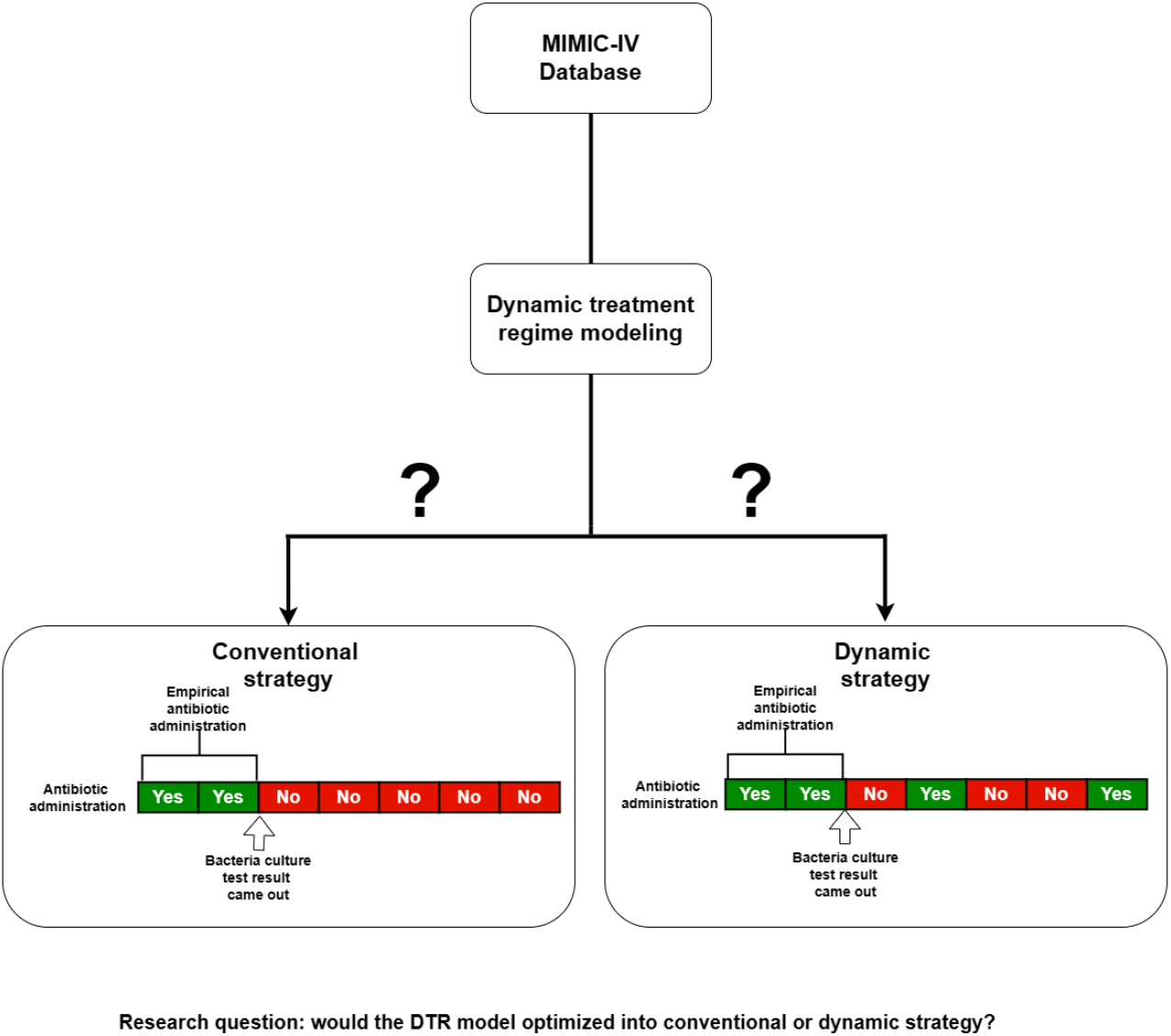
State of art for high risk resistance antibiotics administration strategy optimization research question.

In this study, we would conduct a DTR optimal model by using DWSurv method to allocate the optimal strategy for high risk resisted antibiotics in critical ill patients, to see if the conventional strategy is the optimal one.

## Method

### Data source

This was a retrospective observational cohort study based on MIMIC–IV v2.0[13], a large and freely available database with considerable and high-quality data from patients admitted to an academic tertiary care hospital in the United States between 2008 and 2019. The database included a total of 76,540 intensive care unit (ICU) admissions of 53,150 patients. The author was given access to the database after completing the Collaborative Institutional Training Initiative Program (Record ID Zhaohui Bai) approved by Institutional Review Boards in Beijing TsingHua Changgung Hopistal (waived the requirement for informed consent due to its retrospective nature). The study was designed and implemented in accordance with all relevant guidelines and regulations. The need for informed consent from participants were waived due to the retrospective design of this study.

### Study populations

Adult patients (age ≥ 18 years) who administrated any antibiotics, no matter class or dose, in ICU with drug sensitivity test with “intermediate” or “resistance” were screened for inclusion, and those who didn’t have age or gender were excluded. The intermediate level was defined as C_max_/MIC (minimal inhibitory concentration) is in 1-2, resistance level was defined as C_max_/MIC<1. For those who showed multiple drug resistance (n=3), the administrated antibiotic (none of these patients administrated multiple resisted antibiotics) would be included in study. The index date was set as the first day administrated the resisted antibiotics in ICU.

### Covariates

Data extraction was performed using SQL server. The following static variables (day 0) were extracted from the MIMIC–IV database at the index date: age, sex, Charlson comorbidity index, SOFA, acute physiology score III (APS III), comorbidities. Also, vital signs as dynamic variables (day 1, 2, 3, 4, 5, 6, 14, 28) were collected. We used the maximum or minimum value (or both of them for PCO_2_) of the vital signs and scores depending on which one was worst at each day. Multiple organ dysfunction was assessed using the SOFA and APS III scores. We only included those <20% missingness covariates. The exposure was defined of resisted antibiotics administration for each day, no matter the dose or type. Outcome was 28-day mortality after index date. Detailed definition for variables as shown in (https://github.com/MIT-LCP/mimic-code/tree/main/mimic-iv/concepts).

### Outcome

The main outcome is the 28-day mortality for DTR model. To investigate if model recommendation superior to clinical practice, we collected times that clinical practice obeys or not obey recommendation (obey = 1, not obey = 0) and adding them together to see if the sum of obey is associated with higher 28-day survival.

### Statistical analysis

#### DTR model

We conducted DTR analysis by using R package DWSurv [14], the model recommendation illustration shown in **Supplemental material Table S1**. In brief, the DTR model based on Q-learning estimated a sequence of treatment strategy that maximized the survival time across stages of clinical intervention, the recursive regression would start backward. To locate the censor issue in survival analysis, an inverse probability censoring weighting (IPCW) would be conducted at this stage to fit for survival problems. And recursive optimization would be conducted till first stage. The detailed methodology as shown in **Supplemental material Method**.

#### Model test

To test if the model had any privilege among clinical practice, we added the times of model disobey together (as continuous variable) in day 2-7 as exposure (since too few antibiotic treatments shown in day 14 and day 28), and conducting Cox-regression survival analysis, adjusted with all baseline covariates, to see if disobey times associated with 28-day mortality.

## Result

In total, we included 2,532 ICU admissions with high risk of antibiotic resistance, 652 (25.7%) of them died in 28 days, mean follow-up time was 16.4 days and in total 41,565 person-days. At day 2, 87.4% of them administrated antibiotics, this rate goes down along with treatment process till day 28 (2.1%). The mean age was 63±6 years old, 56.9% of them were male, 69% of them were diagnosed as sepsis according to sepsis 3 criteria, Charlson index median was 5[IQR:4-7]. The baseline characteristics were shown in **Table 1**: inter-quantile range

**Table 1.**
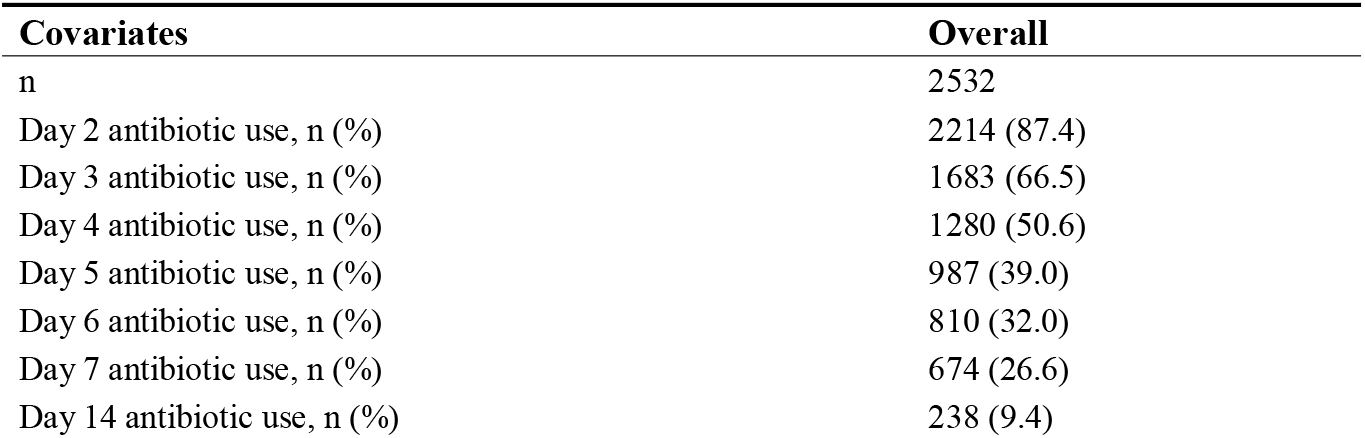
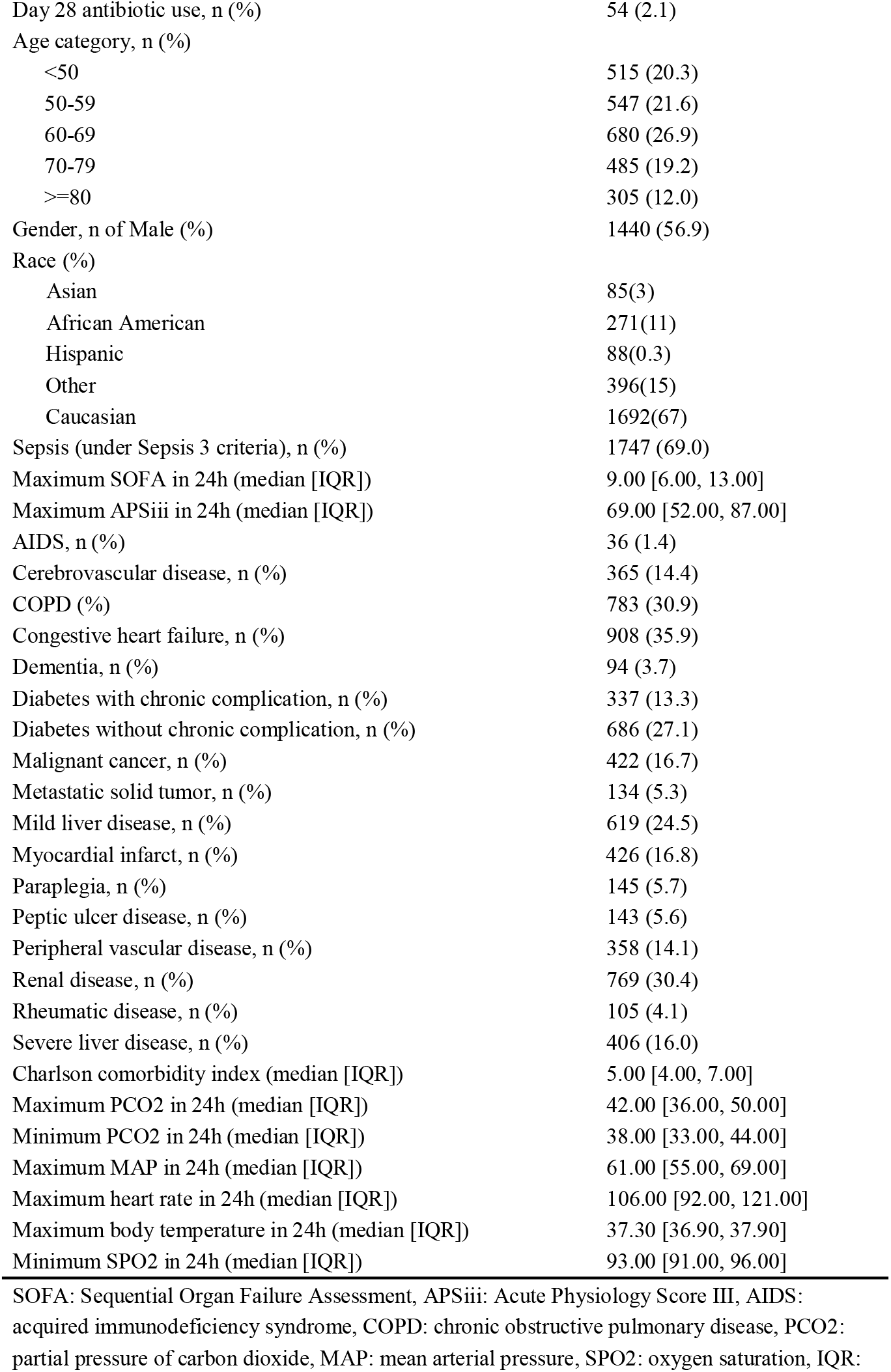
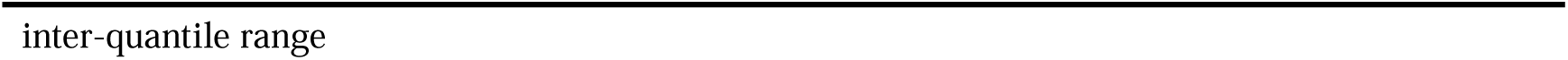
Baseline characteristics for included patients.

| Covariates | Overall |
| --- | --- |
| n | 2532 |
| Day 2 antibiotic use, n (%) | 2214 (87.4) |
| Day 3 antibiotic use, n (%) | 1683 (66.5) |
| Day 4 antibiotic use, n (%) | 1280 (50.6) |
| Day 5 antibiotic use, n (%) | 987 (39.0) |
| Day 6 antibiotic use, n (%) | 810 (32.0) |
| Day 7 antibiotic use, n (%) | 674 (26.6) |
| Day 14 antibiotic use, n (%) | 238 (9.4) |
| Day 28 antibiotic use, n (%) | 54 (2.1) |
| Age category, n (%) |  |
| <50 | 515 (20.3) |
| 50-59 | 547 (21.6) |
| 60-69 | 680 (26.9) |
| 70-79 | 485 (19.2) |
| >=80 | 305 (12.0) |
| Gender, n of Male (%) | 1440 (56.9) |
| Race (%) |  |
| Asian | 85(3) |
| African American | 271(11) |
| Hispanic | 88(0.3) |
| Other | 396(15) |
| Caucasian | 1692(67) |
| Sepsis (under Sepsis 3 criteria), n (%) | 1747 (69.0) |
| Maximum SOFA in 24h (median [IQR]) | 9.00 [6.00, 13.00] |
| Maximum APSiii in 24h (median [IQR]) | 69.00 [52.00, 87.00] |
| AIDS, n (%) | 36 (1.4) |
| Cerebrovascular disease, n (%) | 365 (14.4) |
| COPD (%) | 783 (30.9) |
| Congestive heart failure, n (%) | 908 (35.9) |
| Dementia, n (%) | 94 (3.7) |
| Diabetes with chronic complication, n (%) | 337 (13.3) |
| Diabetes without chronic complication, n (%) | 686 (27.1) |
| Malignant cancer, n (%) | 422 (16.7) |
| Metastatic solid tumor, n (%) | 134 (5.3) |
| Mild liver disease, n (%) | 619 (24.5) |
| Myocardial infarct, n (%) | 426 (16.8) |
| Paraplegia, n (%) | 145 (5.7) |
| Peptic ulcer disease, n (%) | 143 (5.6) |
| Peripheral vascular disease, n (%) | 358 (14.1) |
| Renal disease, n (%) | 769 (30.4) |
| Rheumatic disease, n (%) | 105 (4.1) |
| Severe liver disease, n (%) | 406 (16.0) |
| Charlson comorbidity index (median [IQR]) | 5.00 [4.00, 7.00] |
| Maximum PCO2 in 24h (median [IQR]) | 42.00 [36.00, 50.00] |
| Minimum PCO2 in 24h (median [IQR]) | 38.00 [33.00, 44.00] |
| Maximum MAP in 24h (median [IQR]) | 61.00 [55.00, 69.00] |
| Maximum heart rate in 24h (median [IQR]) | 106.00 [92.00, 121.00] |
| Maximum body temperature in 24h (median [IQR]) | 37.30 [36.90, 37.90] |
| Minimum SPO2 in 24h (median [IQR]) | 93.00 [91.00, 96.00] |

SOFA: Sequential Organ Failure Assessment, APSiii: Acute Physiology Score III, AIDS: acquired immunodeficiency syndrome, COPD: chronic obstructive pulmonary disease, PCO2: partial pressure of carbon dioxide, MAP: mean arterial pressure, SPO2: oxygen saturation, IQR:
inter-quantile range

The detailed DTR optimal model shown as **Supplemental material result parameter**, generally speaking, its suggestion was largely different from clinical practice, as shown in **Figure 2**. In clinical practice, high-risk resistance antibiotic administration fell dramatically along treatment process, since most antibiotics initiation in ICU were based on experience, and the sensitivity result took roughly 3 days after initiation, large amount of antibiotic was stopped at day 3 or 4. On the contrary, the model suggested treatment with gaps in 7 days, rather than a continuous de-escalation pattern.

**Figure 2.**
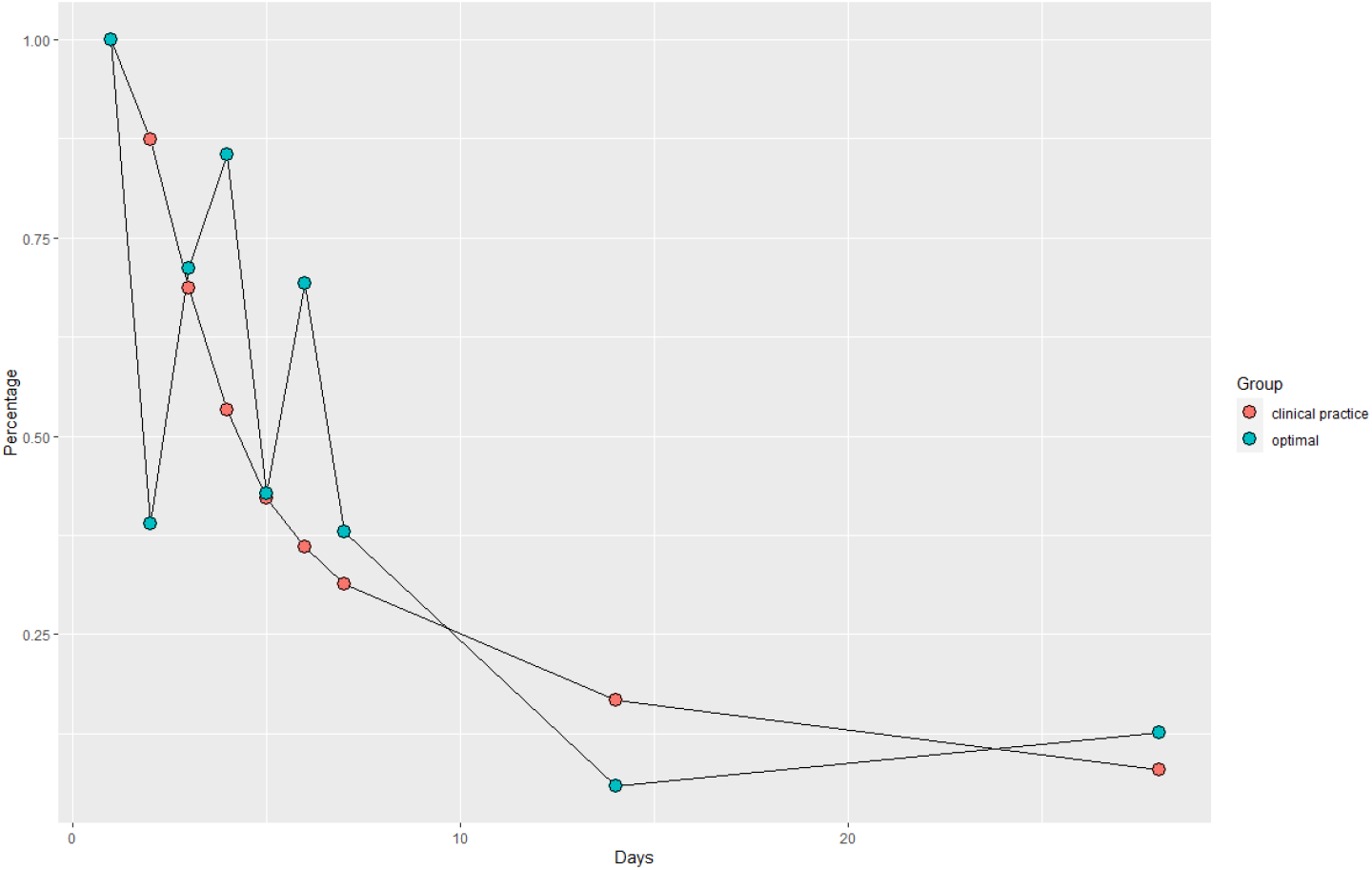
DTR model optimal suggestion and clinical practice for antibiotic administration for each day.

In model test, the crude 28-day mortality showed a clear dose-response to disobey times, the mortality rate was 15%, 19%, 22%, 25%, 29%, 31%, 36% for 0-6 disobey times, respectively, as shown in **Table 2**:

**Table 2.** 28-day mortality for clinical practice disobey model times.

| Disobey times | Alive | Death | Mortality rate |
| --- | --- | --- | --- |
| 0 | 11 | 2 | 15% |
| 1 | 110 | 26 | 19% |
| 2 | 480 | 137 | 22% |
| 3 | 699 | 237 | 25% |
| 4 | 426 | 179 | 29% |
| 5 | 136 | 61 | 31% |
| 6 | 18 | 10 | 36% |

Also, we conducted Cox-regression for survival analysis among model obedience, adjusted by all baseline covariates listed in **Table 1**, the result showed a significant association between disobey times and mortality, the adjusted hazard ratio for disobey times is HR=1.21 [95%CI:1.12-1.30], P<0.001.

## Discussion

In this study, we found out that DTR model showed the optimal high-risk resistance antibiotic administration strategy was a dynamic strategy with pauses, rather than a de-escalation strategy, or simply stop using certain antibiotic when high-risk of resistance was found out. The survival analysis showed this model would have lower mortality rate than clinical practice, since the disobey times were highly correlated to 28-day mortality HR=1.21 [95%CI:1.12-1.30]. Specifically, in theory, the “competition-maximizing” strategy can contain mixed populations of sensitive and resistant bacterial below a threshold density for significantly longer than matched populations containing only resistant bacterial (PMID: 32413038). Although we can’t observe the bacteria competition directly, we proved this theory, at least in part, is feasible in clinical practice with a proper DTR model. To our knowledge, this is the first study allocating this issue among critical ill patients. It’s need to clarify that no one should interpret our result as “clinician need to stop using resisted for 1-2 days and restarted again once MIC result found it resisted”. This study is about to investigate a potential new perspective in personalized antibiotics administration, however, it is still vague for clinical practice.

On the other hand, large number of clinicians would restart resisted antibiotics in clinical practice, which usually would be considered as “miss used” in conventional way. For example, in MIMIC-IV database a handful ICU doctors did restart them (perhaps due to patients worsen condition or lack of available treatment), otherwise we won’t able to optimize a DTR model if they didn’t use it in an extraordinary way.

There were some evidences suggested that DTR model is better than clinical practice, one study found out that a DTR strategy based on reinforcement learning is

## Data Availability

All data produced in the present study are available upon reasonable request to the authors

## Supplemental Material

**Supplementary Table S1.**
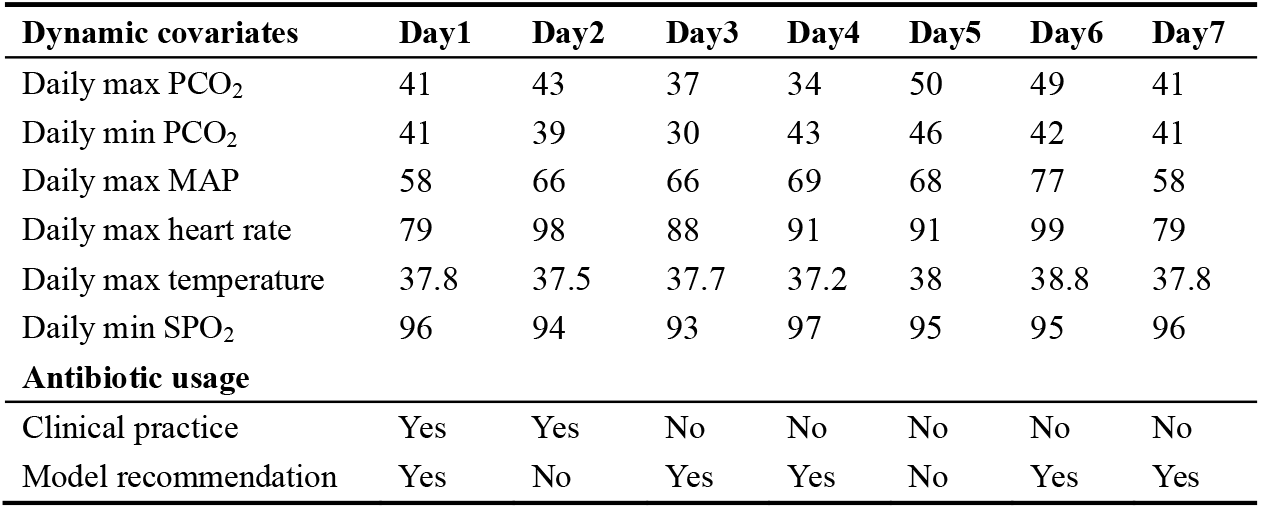
Pseudo-patient (age=73, Charlson Index=4, gender=male, first 24h max SOFA=11, first 24h max APSiii=92) resisted antibiotic administration in clinical practice and dynamic treatment regime recommendation.

### Supplemental Method

The general idea of this study is shown as **Figure S2**:

We defined 8 stages at day 2-7,14 and 28 after antibiotics administration and recorded the survival times within each stage. The DTR model used a backward induction algorithm to estimate the sequence of optimal rules. The decision stage started at day 2 since every patient had been administrated at day 1 (e.g. day 2 means stage 1, day 3 means stage 2 … day 28 means stage 8, etc.). We define the survival time *T*_*i*1_ for patient *i* = 1,2, …, *n* corresponded to the time spent in the first stage, and *η*_*i*2_ denotes to a random variable that indicates if patient *i* enters the second stage and *T*_*i*2_ is the time from the beginning of the second stage to the event and is defined only for patients who enter the second stage (*η*_*i*2_ = 1), so that the survival time at stage 2 would be: *T*_i1_ + *η*_*i*2_*T*_*i*2_ and the survival times for other stages were defined similarly.

1. In the first step, the optimal stage 8 decision rule (day 28) was estimated by modeling the counterfactual accelerated failure survival time in stage 8 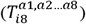 as a function of the treatment received on day 28 (no antibiotic [*a*_8_ = 0] or with antibiotic [*a*_8_ = 1] and of the feature variables measured on day 28 or before (*h*_8*β*_ and *h*_8*ψ*_):

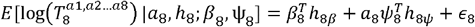

The term 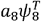 is the stage 8 blip function. It represents the effect of antibiotics instead of without antibiotics and its interaction with feature variables. The term *ψ*_8_ is a vector of coefficients for feature variables and *ψ*_8*ψ*_ represents information (previous treatment, covariates and survival times) available prior to making the stage 8 treatment.
2. Propose models for the probability of treatment at this stage *P*[*A*_*i*8_ = 1|*h*_*i*8,_*η*_*i*8_ = 1;*α*_8_] (treatment model), and censoring probability: *P*[Δ_*i*8_ = 1|*h*_*i*8_,*η*_*i*8_ = 1;*λ*_8_] (censor model). Fit the two models using subjects who enter 8^th^ stage (*η*_8_ = 1), in our case, using logistic regression.
3. Estimate the parameters (*β*_8_,ψ_8_) by solving the weighted estimating equations:

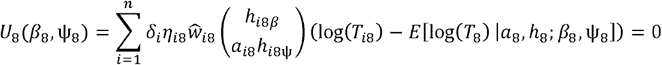

Under weights that satisfy a balancing property, as inverse-probability weighting:

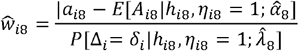

The optimal stage 8 treatment was identified for each subject who entered stage 8 by 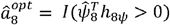. If optimal rule recommends antibiotic administration on day 28 if the condition 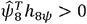 is satisfied, and no antibiotic administration otherwise.

Secondly, we started the recursive step, constructing the counterfactual survival time 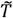. It represents the survival time from day 14 (stage 7) onwards had the patient received his optimal stage 8 treatment. There were 3 possibilities for this variable:

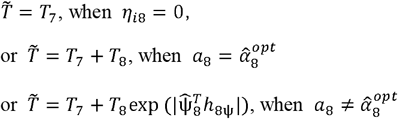

By using this pseudo-survival time as outcome, the treatment decision optimization at stage 7 would follow the same optimal rule, repeat this recursive step till stage 1.

### Supplemental Result parameter

#Stage 1: treat if 0.2867 - 0.0028 pco2_max1 + 0.0056 pco2_min1 - 0.0012 map1 - 0.0006 hr1 + 0.0270 temp1 - 0.0085 spo21 + 0.0912 age1 - 0.0072 sofa1 - 0.0066 apsiii1 - 0.0282 charlson1 - 0.2227 gender1 > 0

#Stage 2: treat if 2.3503 + 0.0009 pco2_max2 + 0.0058 pco2_min2 - 0.0064 map2 + 0.0005 hr2 - 0.0739 temp2 + 0.0019 spo22 - 0.0637 age2 - 0.0212 sofa2 + 0.0095 apsiii2 + 0.0373 charlson2 - 0.0263 gender2 > 0

#Stage 3: treat if 2.3846 + 0.0011 pco2_max3 - 0.0045 pco2_min3 - 0.0102 map3 + 0.0016 hr3 - 0.0299 temp3 - 0.0050 spo23 + 0.0059 age3 + 0.0017 sofa3 - 0.0013 apsiii3 - 0.0055 charlson3 + 0.1561 gender3 > 0

#Stage 4: treat if -0.6302 + 0.0010 pco2_max4 - 0.0028 pco2_min4 + 0.0047 map4 - 0.0028 hr4 + 0.0359 temp4 - 0.0077 spo24 - 0.0410 age4 + 0.0075 sofa4 - 0.0008 apsiii4 + 0.0305 charlson4 - 0.0633 gender4 > 0

#Stage 5: treat if 0.2246 - 0.0002 pco2_max5 + 0.0069 pco2_min5 - 0.0020 map5 + 0.0008 hr5 - 0.0107 temp5 - 0.0011 spo25 + 0.1175 age5 + 0.0026 sofa5 + 0.0001 apsiii5 - 0.0478 charlson5 + 0.0486 gender5 > 0

#Stage 6: treat if 0.2098 - 0.0024 pco2_max6 + 0.0002 pco2_min6 + 0.0007 map6 - 0.0023 hr6 + 0.0374 temp6 - 0.0140 spo26 - 0.0292 age6 - 0.0119 sofa6 + 0.0004 apsiii6 + 0.0232 charlson6 + 0.0010 gender6 > 0

#Stage 7: treat if 2.1468 + 0.0037 pco2_max7 - 0.0027 pco2_min7 + 0.0002 map7 - 0.0002 hr7 - 0.0366 temp7 - 0.0060 spo27 - 0.0260 age7 + 0.0009 sofa7 - 0.0012 apsiii7 + 0.0120 charlson7 - 0.0466 gender7 > 0

#Stage 8: treat if 0.0660 - 0.0007 pco2_max14 - 0.0012 pco2_min14 - 0.0019 map14 + 0.0015 hr14 + 0.0408 temp14 - 0.0120 spo214 + 0.0183 age14 - 0.0255 sofa14 + 0.0010 apsiii14 - 0.0081 charlson14 + 0.0461 gender14 > 0

#Stage 9: treat if -0.0493 - 0.0020 pco2_max28 - 0.0005 pco2_min28 - 0.0016 map28 - 0.0008 hr28 + 0.0161 temp28 + 0.0029 spo228 + 0.0101 age28 - 0.0188 sofa28 - 0.0015 apsiii28 + 0.0216 charlson28 + 0.0213 gender28 > 0

